# Driving Factors in Pediatric Emergency Department Use: an Ecological Retrospective Study

**DOI:** 10.1101/2025.08.18.25333899

**Authors:** Denis Mongin, Hervé Spechbach, Joachim Marti, Frederic Ehrler, Johan N. Siebert

## Abstract

**Background:** Pediatric emergency departments (PEDs) often face high volumes of low-acuity visits, reflecting gaps in primary care access and socio-economic disparities. We investigated how neighborhood socio-economic vulnerability, pediatrician availability, and proximity to the PED jointly influence PED utilization in Geneva, Switzerland.

**Methods:** In this retrospective ecological study (Jan 2023-Dec 2024), we aggregated all PED visits for children aged 0-16 years by neighborhood and Canadian Triage Acuity Scale (CTAS) level. Neighborhood visit incidence (unique patients per child population) was modeled using mixed-effects regression against a composite socio-economic vulnerability index (NSVI), pediatrician density within a 2 km radius, and distance to the PED, incorporating an exponential decay function for distance and postal code as a random intercept.

**Results:** There were 68,482 PED visits by 35,994 children (35.1% of Geneva under-16 population). Low-acuity visits (CTAS 4-5) comprised ∼50% of encounters. Both distance and socio-economic vulnerability showed clear dose-response relationships, with stronger effects observed for lower-acuity visits, and no interaction effect between them. Overall, proximity accounted for up to 20.8% of non-urgent PED use, while neighborhood socio-economic vulnerability explained up to 19.7% of low acuity visits across Geneva. Pediatrician density showed a modest inverse association for low-acuity visits only.

**Conclusions:** Both proximity and socio-economic vulnerability are independent determinants of non-urgent PED use. Policies focusing only on primary care access risk missing key drivers of PED use, highlighting the need for locally tailored strategies such as community outreach near hospitals or programs to strengthen health literacy among families.

**Summary:** Proximity and socio-economic vulnerability each explain up to 20% of non-urgent pediatric emergency visits, highlighting major contextual drivers of avoidable care use.

**What’s Known on This Subject:** Pediatric emergency department crowding, often driven by low-acuity visits, has been linked to socio-economic disadvantage, poor primary care access, and proximity. Yet, few studies have jointly examined these factors or compared their impact within a universal healthcare setting.

**What This Study Adds:** This study disentangles the effects of distance, socio-economic vulnerability, and pediatrician density on PED use. Proximity and disadvantage each explain ∼20% of low-acuity visits, while pediatrician density has limited impact, highlighting the need for targeted, locally adapted interventions.

## 1 BACKGROUND

Pediatric emergency departments (PEDs) are essential components of healthcare systems, ensuring 24/7 access to acute care for children. However, high utilization raises significant public health concerns, often reflecting gaps in primary care access, parental health literacy, and the relative convenience of emergency services[1–4]. In the United States, children under 15 account for a substantial share of emergency department (ED) visits, with over 26 million annual PED visits reported[5]. An estimated 54 to 87% of these visits are attributed to non-urgent conditions[6, 7]. Similar trends have been documented across European healthcare systems[1, 8], where increased PED attendance contributes to overcrowding, extended waiting times, operational inefficiencies, and a higher incidence of patients leaving without being seen [9, 10]. These factors can lead to reduced patient satisfaction and a deterioration in the overall quality of emergency care.

A robust body of literature indicates that PED utilization is strongly influenced by socio-economic determinants. Children from lower socio-economic status (SES) families and those covered by public insurance consistently exhibit higher rates of ED use compared to their more affluent or privately insured counterparts[11]. In addition to socio-economic vulnerabilities, ethnicity and language also play influential roles in shaping patterns of PED use. Data from North America indicate that children from minority background are more likely to visit the ED, with disparities reflecting deeper systemic inequities such as poverty, limited health literacy, linguistic barriers, and the absence of a consistent medical home[11].

In addition to socio-economic determinants, access to primary care has also been proven to influence PED attendance[12]. Notably, studies have shown that enhancing access to high-quality primary care is associated with significant reductions in ED visits for low-acuity conditions[13].

Beyond individual socio-economic factors, the geographic and ecological context in which families live also significantly influences ED utilization. Geographic proximity to healthcare facilities is a well-established driver of care-seeking behavior[14, 15]. Families living closer to an ED are more likely to use it, even for less urgent conditions[11]. Conversely, those in peripheral areas may delay care due to transportation or access barriers, often presenting with more severe illness.

While distance to PED, socio-economic conditions, and access to primary care are each recognized as key determinants of PED utilization, these factors are often interrelated and may interact in complex ways. Socio-economic disadvantage can limit access to primary care, just as geographic proximity to PED services can be shaped by underlying social and spatial inequalities. Moreover, regions with poor access to pediatricians often overlap with socio-economically deprived areas[16, 17], further compounding barriers to appropriate care. Despite these interconnected dynamics, most existing studies have examined these determinants in isolation, without accounting for their combined or interactive effects. This fragmented approach may obscure important residual confounding, limiting our understanding of how these factors jointly influence PED utilization. Furthermore, comparing the relative impact of each of these factors is essential to identify the most influential drivers of PED use and to inform more targeted, effective interventions.

To address these gaps, this retrospective ecological study systematically examined the demographic, socio-economic, the density of pediatricians and spatial profiles of children presenting to a tertiary PED over a two-year period. Our aim was to analyze patterns of PED use in relation to the availability of primary care, socio-economic background, and geographic proximity to the PED, in order to better understand the relative contribution of these factors.

## 2 METHODS

### 2.1 Study design

We conducted a retrospective ecological study of PED visits to Geneva’s tertiary hospital over a two-year period. Geneva is divided into 476 neighborhoods (median population: 693; median area: 0.33 km²), grouped into 65 postal zones. Neighborhood-level PED utilization among children aged 0–16 was analyzed by triage acuity using the five-level Canadian Triage and Acuity Scale (CTAS: 1=resuscitation, 2=emergent, 3=urgent, 4=less urgent, 5=non-urgent)[18]. Due to low numbers, CTAS levels 4 and 5 were combined (hereafter referred to as level 4*). Neighborhoods with fewer than 10 children were excluded.

### 2.2 Setting

Geneva is a densely populated urban canton (530,246 residents, including 86,835 children under 16), with substantial cross-border commuter traffic[19]. The Geneva University Hospitals (HUG) is a 2,095-bed tertiary care institution comprising eight hospital sites and two clinics.

### 2.3 Outcome

The primary outcome was the neighborhood-level incidence of PED use per triage level, defined as the number of unique pediatric patients per 100 children aged 0-16 in the neighborhood population. Patient addresses were geocoded using the state geocoding API[21], and age-specific neighborhood population data were provided by the Geneva State Geomatics Services.

### 2.4 Exposures of interest

The primary exposures were distance to the PED, neighborhood socio-economic status, and pediatrician density. Distance to PED was measured from the geographic centroid of each neighborhood to the PED. Socio-economic Status was assessed using the Neighborhood Socio-economic Vulnerability Index (NSVI), a composite score ranging from 0 to 6 state-provided indicators, including income, unemployment, and social benefits (full criteria are provided in Supplementary Table 1). Pediatrician density was calculated as the number of non-hospital pediatricians per 1,000 children within a 2 km radius of each neighborhood centroid, using data from the Swiss Federal Register of Medical Professions.

### 2.5 Statistical analysis

#### 2.5.1 Main analysis

We modeled neighborhood PED incidence by acuity level with a nonlinear mixed-effects regression:

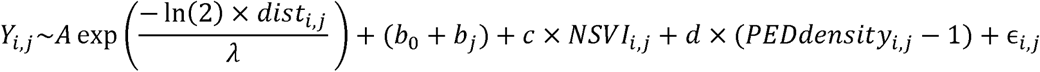

With indexing neighborhoods and *i* indexing postal codes,

- *Y_i,j_*: incidence of PED use in neighborhoods and postal code %
- *A* Maximum distance-related increase in PED incidence
- λ the halving distance (distance at which the distance effect reduces by half)
- *c, d* : Effects of NSVI and pediatrician density, respectively
- *b*_0_: baseline PED use incidence for a wealthy neighborhood (NSVI = 0), pediatrician density of 1 per 1000 children, and zero distance effect.
- *B_j_*: random intercept by postal code

Absence of spatial correlation in the regression residuals was assessed using Moran’s Index[20]. Quality of the regression was assessed using the Efron pseudo R-squared[21], which ranges from 0 to 1, with 1 indicating a perfect fit. All spatial statistical analyses were conducted using R[23], *data.table* for data management, and *sp* and *sf*[24] for handling spatial data.

#### 2.5.2 Sensitivity analysis

We stratified the main model by age groups (0–5 vs. 6–15 years) and time of visit, distinguishing between school hours (7 AM-6 PM), non-school hours (7 PM-6 AM), and weekends. Additionally, we tested for an interaction between the NSVI and distance using an extended model that included an interaction term for the non-linear exponential effect of distance and NSVI (see Supplementary Material for full model specification).

#### 2.5.3 Counterfactual analyses

To quantify potential reductions in PED use, we simulated four scenarios using the adjusted model and Geneva: placing all neighbourhoods at a fixed distance of 5km from the PED; setting all neighbourhoods to the lowest NSVI (i.e., 0); assuming a uniform pediatrician density of 2 per 1,000 children; and combining all three conditions simultaneously.

### 2.6 Ethical consideration

The study was exempt from formal ethical review under the Swiss Human Research Act, as confirmed by the Geneva Research Ethics Commission (req-2025-00269), and was approved by HUG’s institutional board for non-HRA research.

## 3 RESULTS

### 3.1 Demographics

Between January 1, 2023, and December 31, 2024, the PED recorded 68,482 visits by 35,994 unique patients, representing 35.1% of the pediatric population. CTAS level 1 (highest acuity) accounted for 2.1% of visits, mainly for respiratory issues. Level 2 comprised 11.3%, mostly respiratory and infectious cases. Level 3 represented 36.2%, with common complaints including respiratory, gastrointestinal, and musculoskeletal conditions. Level 4* was the largest group (50.5%), primarily involving infectious, gastrointestinal, musculoskeletal, dermatologic, ear, nose and throat (ENT), and ophthalmologic presentations.

Higher acuity was associated with younger age (median 3.5 years at level 1 vs. >5 years at level 4*), a higher proportion of male patients, fewer residents of Geneva (dropping from 88.9% at level 4* to 81.9% at level 1), and a greater proportion of Swiss nationals and French speakers (Table 1). Low-acuity visits peaked between 7 and 10 AM (up to 55%) and increased by 10 percentage points during the summer months. The age distribution showed an inverse U-shape, peaking at around 60% among children aged 4– 8 and declining to 35% for both younger and older children (Supplementary Figure 1).

**Table 1:**
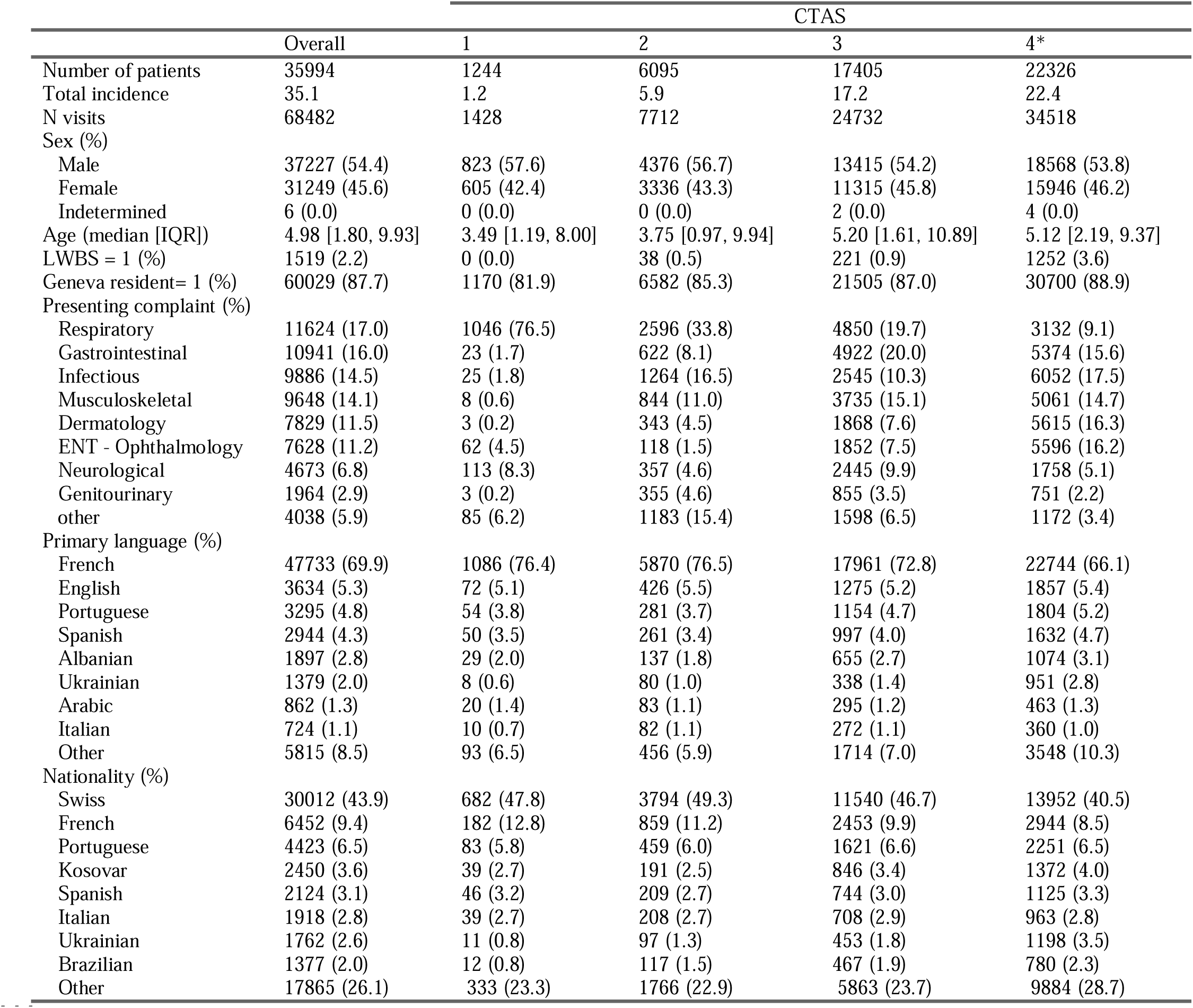
Descriptive statistics and demographic characteristics of PED visits between January 1, 2023, to December 31, 2024 at Geneva University Hospitals, for all acuity triage level, and per Canadian Triage Acuity Scale (CTAS) level (1 : vital resuscitation, 2 : emergent, 3 : urgent, 4* : less urgent (4) and non-urgent (5)).

### 3.2 Neighborhoods aggregated statistics

#### 3.2.1 Pediatrician and socio-economic indicators

A total of 120 private pediatricians operated across 67 practices, yielding a crude density of 1.6 per 1,000 children (vs. 1.2 nationally)[22]. Pediatrician density varied substantially across the neighborhoods, with a median of 1.32 [0.80, 1.97] pediatricians per 1000 children. The study included 357 neighborhoods, with a median pediatric population of 183 [IQR 79–330] and a median distance of 3.84 km [IQR 2.29–6.88] from the PED. NSVI scores showed 50.7% of neighborhoods had no vulnerability, while 5.3% met all six NSVI indicators, qualifying as highly vulnerable. Distributions of key indicators are visualized in Supplementary Figure 2.

#### 3.2.2 PED statistics

The incidence of PED use varied substantially across neighborhoods (Figure 1), with a mean neighborhood-level incidence of 33.9% (SD 16.0%) across all triage levels. In the multivariable analysis, significant spatial dependence of PED utilization was observed for CTAS levels 2, 3, and 4*, with the strength of this dependence increasing as triage acuity decreased (Table 2). For CTAS 2, the mean neighborhood-level incidence was 5.8%, with a distance-related variation of 4.8 percentage points [95% CI: 2.9-6.7], representing 82% of the mean incidence. The spatial effect decreased by half approximately every 0.85 km [0.31-1.4]. For CTAS 3, the distance-related variation was 14 percentage points [10–18] (83% of the mean incidence), with a halving distance of 2.8 km [0.74-4.8]. For CTAS 4*, the variation was even larger, at 27 percentage points [22–31], representing 130% of the mean incidence, but with a faster decay over distance, halving every 1.7 km [1.1-2.3]. The spatial extent of the distance effect was similar for CTAS 3 and 4*—approximately 5km—but the amplitude of the effect was markedly greater for CTAS 4* (Figure 2).

**Figure 1:**
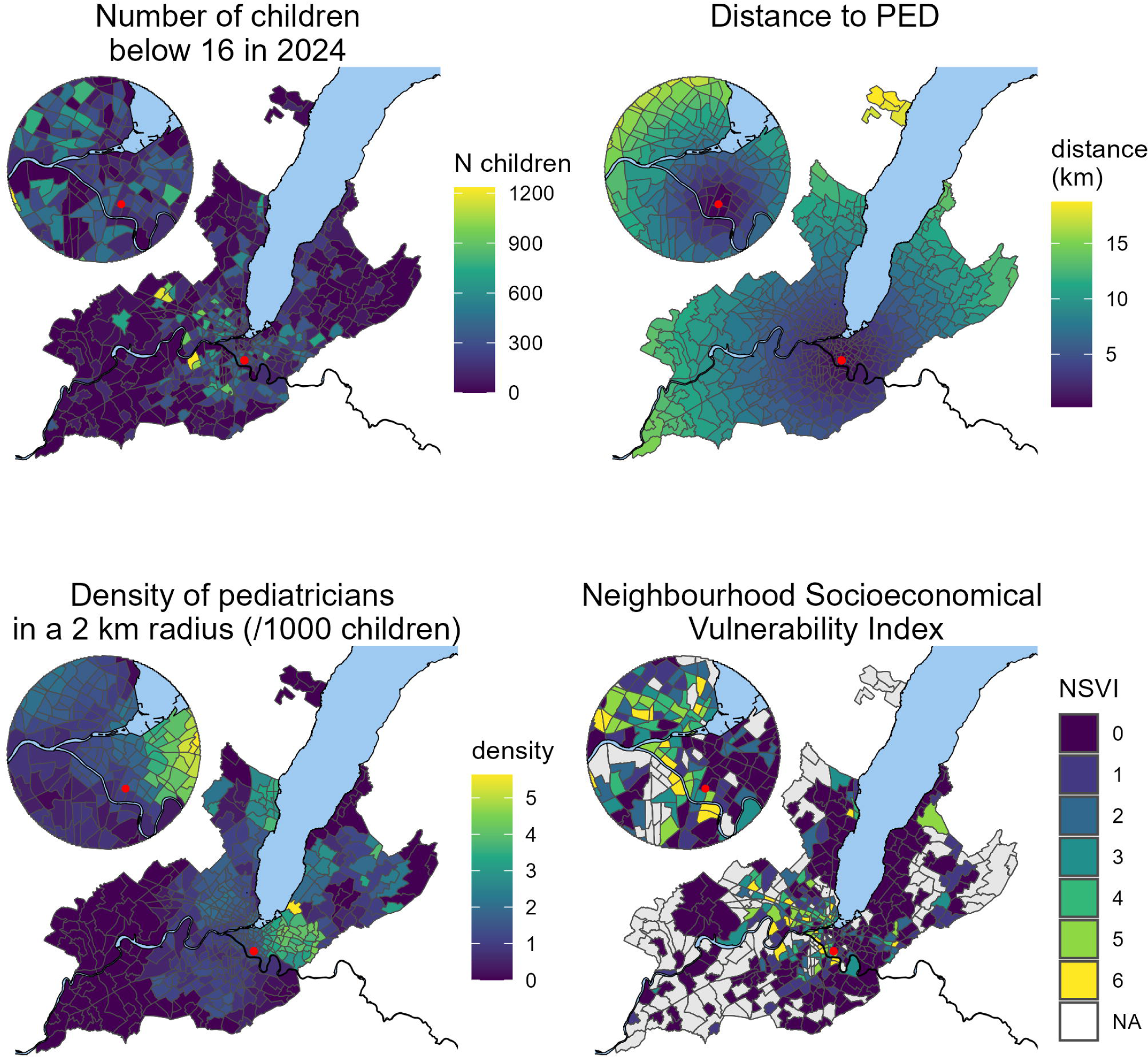
incidence of PED use during 2023-2024 (Number of children going to PED during the period as a percentage of the number of children in the neighborhood), per neighbourhood and Canadian Triage Acuity Scale (CTAS) level (2: emergent, 3: urgent, 4*: less urgent (4) and non-urgent (5)). The Red point is the location of the PED in Geneva.

**Table 2:**
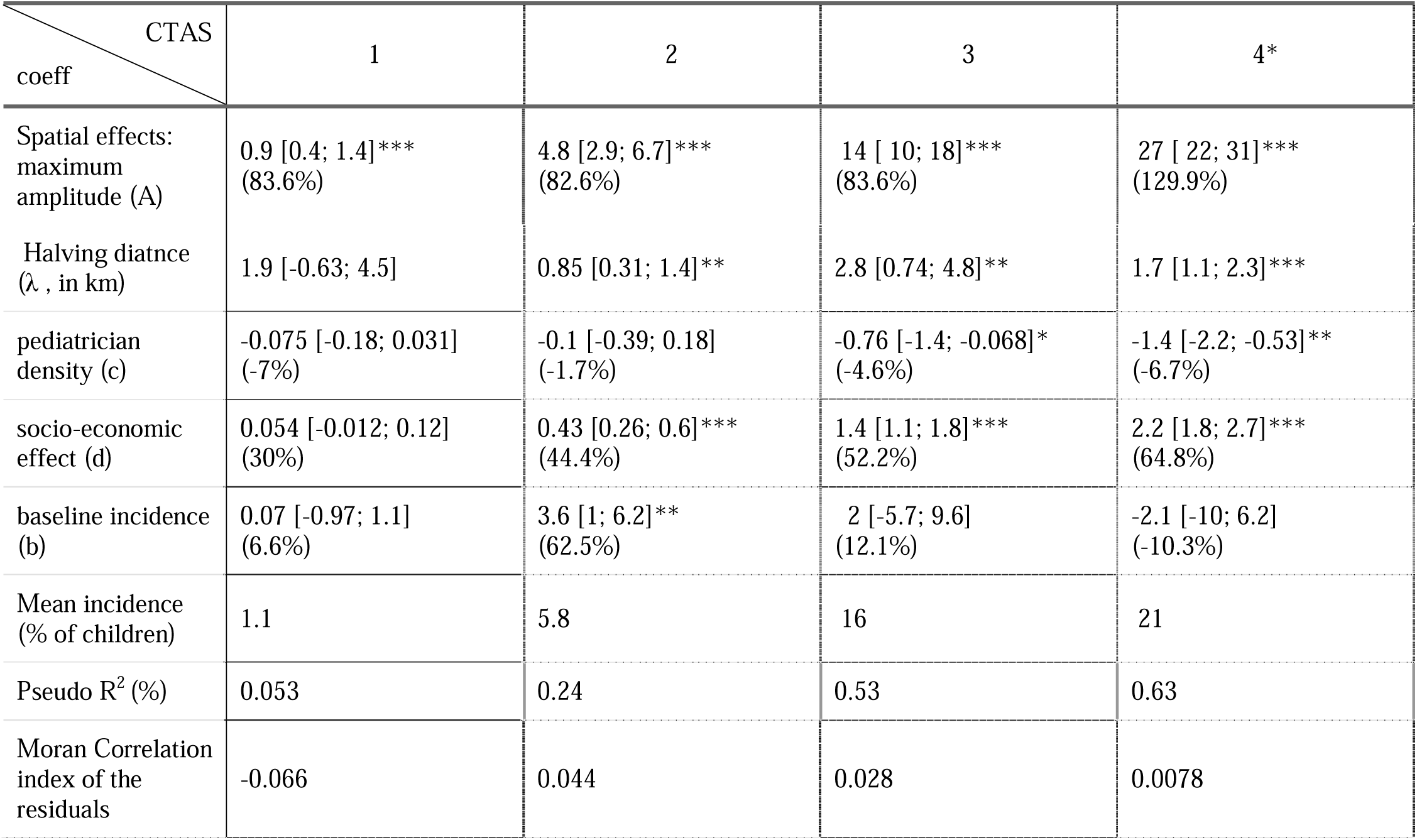
Results of the multivariable regressions modeling the incidence of PED use during 2023-2024 as a function of distance to the PED, neighborhood socio-economic vulnerability, and pediatrician density, stratified by Canadian Triage Acuity Scale (CTAS) level (1 : vital resuscitation, 2 : emergent, 3 : urgent, 4* : less urgent (4) and non-urgent (5)). and represent the amplitude and characteristic length of the exponential spatial decay, respectively. is the coefficient for pediatrician density (expressed as the number of pediatricians per 1000 children), is the coefficient for Neighbourhood Socio-Economic Vulnerability Index (NSVI), and b denotes the baseline incidence. The percentage in parenthesis represents the marginal variation induced by the variable compared to the mean incidence.

**Figure 2:**
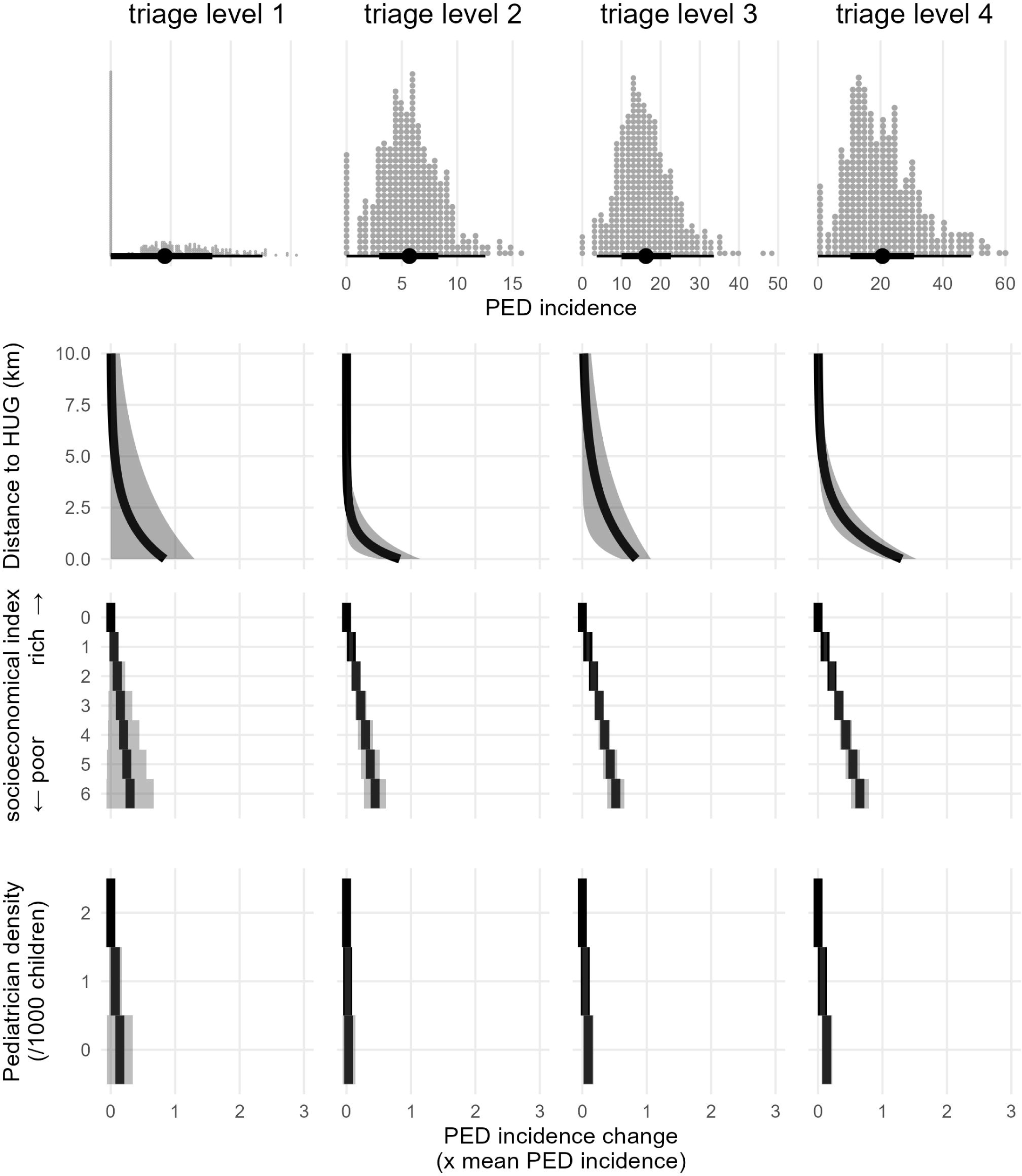
Effects of key exposure variables on PED use, stratified by triage level (columns). The top row shows the distribution of the neighborhood-level PED incidence. The three lower rows display variation in PED use incidence associated with (1) distance to the PED, (2) the neighborhood socio-economic index, and (3) pediatrician density. Effects are expressed relative to the mean incidence for each triage level. HUG, Geneva University Hospitals. PED, Pediatric Emergency Department.

Socio-economic vulnerability was also significantly associated with PED use for CTAS 2–4*, with the effect size increasing as acuity decreased. Each additional NSVI point increased CTAS 2 incidence by 0.43 percentage points [0.26–0.6], totaling a 2.56-point gap (44% of the mean incidence) between least and most vulnerable areas. For CTAS 3 and 4*, these gaps were 8.4 (52%) and 13.2 (64.8%) percentage points, respectively.

Higher pediatrician density was significantly associated with lower PED use for CTAS 3 and 4* only. Each additional pediatrician per 1,000 children reduced incidence by 0.7 [0.07–1.4] and 1.4 [1.1–1.8] percentage points, respectively, corresponding to 4.6% and 6.7% of the mean incidence. Residual spatial autocorrelation (Moran’s I) was below 0.1 and non-significant in all models. Model fit (Efron’s pseudo R²) improved as acuity decreased, reaching a maximum at 0.63 for CTAS 4* (Table 2).

### 3.3 Sensibilty analysis

No interaction was found between distance and NSVI, indicating that the effect of distance was consistent across socio-economic levels (Supplementary Table 1). Subgroup analyses showed that both distance and socio-economic effects were more pronounced among older children (≥6 year) than younger children (≤5 years) (Supplementary Tables 2 and 3; Supplementary Figures 3 and 4). Similarly, all effects were stronger during non-school hours and weekends compared to school hours (Supplementary Tables 4-6; Supplementary Figures 5-7).

### 3.4 Conterfactual approach

Model-based estimates predicted 980, 5,291, 15,231, and 19,474 unique patients for CTAS levels 1 to 4*, respectively, closely matching the observed data (Table 3 vs. Table 1). Counterfactual scenarios estimated reductions in visits under three conditions: 1) setting all neighborhoods at an equal distance of 5 km was predicted to reduce visits by 571 patients (−10.8%) for CTAS 2, 1,829 (−12%) for CTAS 3, and 4,050 (−20.8%) for CTAS 4*; 2) assuming the least vulnerable socio-economic profile reduced visits by 736 (−13.9%) for CTAS 2, 2,448 (−16.1%) for CTAS 3, and 3,832 (−19.7%) for CTAS 4*; and 3) applying a uniform pediatrician density of 2 per 1,000 children resulted in modest reductions of 37 (−0.7%) for CTAS 2, 282 (−1.9%) for CTAS 3, and 516 (−2.6%) patients for CTAS 4*.

**Table 3.** Estimated number of PED visits during the 2023-2024 period, as predicted by the adjusted regression model under real-world conditions and four counterfactual scenarios: (1) all Geneva neighborhoods located 5 km from the PED, (2) all neighborhoods classified as socio-economically wealthy, (3) all neighborhoods with a pediatrician density of 2 per 1,000 children, and (4) all of the above conditions combined. Results are stratified by Canadian Triage Acuity Scale (CTAS) level (2: emergent, 3: urgent, 4*: less urgent (4) and non-urgent (5)) for cases where the regression model provide significant effects.

| CTAS | 2 | 3 | 4* | all |
| --- | --- | --- | --- | --- |
| Counterfactual scenarios |  |  |  |  |
| Full model: Number of predicted patients | 5291 | 15231 | 19479 | 40981 |
| Distance to PED = 5km:<br>Number of patients<br>(difference with full model: N, %) | 4720<br>(-571, -10.8%) | 13402<br>(-1829, -12%) | 15429<br>(-4050, -20.8%) | 34531<br>(-6450, -15.7%) |
| NSVI = 0 (wealthy):<br>Number of patients<br>(difference with full model: N, %) | 4555<br>(-736, -13.9%) | 12783<br>(-2448, -16.1%) | 15647<br>(-3832, -19.7%) | 33873<br>(-7108, -17.3%) |
| Pediatrician density = 2 per 1,000 children<br>Number of patients<br>(difference with full model: N, %) |  | 14949<br>(-282, -1.9%) | 18963<br>(-516, -2.6%) | 40118<br>(-863, -2.1%) |
| All counterfactual combined<br>Number of patients<br>(difference with full model: N, %) | 3932<br>(-1344, -25.5%) | 10634<br>(-4557, -30%) | 11029<br>(-8365, -43.1%) | 26452<br>(-14386, -35.2%) |

## 4 DISCUSSION

### 4.1 Main findings and Interpretation

This retrospective study revealed substantial variation in neighborhood-level PED use among children aged 0-16 across Geneva, a high-income city with universal health coverage and advanced medical infrastructure. High acuity visit patterns were relatively uniform across neighborhoods, suggesting that severe emergencies are less affected by contextual factors. In contrast, lower-acuity visits showed marked spatial variability, with a sharp exponential increase in PED utilization within a 5 km radius of the hospital. This distance effect varied with triage acuity, from no significant impact for high-acuity cases to contributing up to 20% of visits for the lowest-acuity cases. Socio-economic vulnerability, independent of distance, was also strongly associated with higher PED use, with effects becoming more pronounced at lower acuity levels and accounting for up to 20% of visits for triage level 4. Pediatrician density was associated with PED use only for CTAS levels 3 and 4, and its overall effect was modest, accounting for less than 2% of visits.

#### 4.1.1 Geographic Proximity

Proximity was a key determinant of PED use. Children living within 5 km of the hospital were more likely to visit, especially for non-urgent conditions. This pattern reflects the well-documented “distance decay effect”, whereby even modest travel distances reduce the likelihood of seeking care for less urgent needs[4, 11, 15]. Previous studies have reported mixed findings, likely due to methodological differences, as many modeled distance linearly or with simple thresholds [15]. By using an exponential decay model, we captured the sharp, short-range effect of distance accurately, which is particularly relevant in a dense urban environment like Geneva. Although some studies have evidenced stronger distance effects in socio-economically disadvantaged areas [23], we observed no significant interaction between distance and socio-economic vulnerability, suggesting that the effect of distance was consistent across the social gradient. Our findings align with Ludwick et al. [24], who similarly reported distance decay in PED use, with incidence halving every 2-5 km depending on acuity—an effect we found most prominent for CTAS level 4*. This reinforces the notion that even in compact, well-connected cities, spatial barriers (e.g., crossing city or canton boundaries) can meaningfully influence care-seeking behavior, particularly for non-urgent cases.

#### 4.1.2 Socio-Economic Vulnerability

Socio-economic vulnerability was a strong, independent predictor of PED use, with an effect size comparable to that of distance. Neighborhoods with higher composite vulnerability scores had significantly higher visit rates, particularly for lowest acuity cases. For CTAS levels 4 and 5, the incidence gap between the most and least vulnerable areas reached 13.2 percentage points (about 65% of the mean incidence) and accounted for up to 20% of total visits. This pattern is consistent with findings from other high-income countries, such as the UK and US, where lower-income communities disproportionately rely on emergency services for conditions that may not require hospital-level care[11, 23, 25, 26]. Our findings are in line with a previous Swiss multicenter study, which showed that families facing financial hardship had roughly 2–3 times higher odds of using PED services for non-urgent conditions[6]. This supports the view that low-acuity emergency visits are more strongly linked to financial insecurity than to factors such as parental education, employment status, immigrant background, or lack of a support network. Other known contributors[4] include limited system or health literacy[27–29], language or cultural barriers[29], lack of a consistent primary care physician[30, 31], and the need for parental reassurance[32]. For newly arrived or non-native families, the PED is often perceived as a trustworthy first point of contact when navigating uncertain or unfamiliar health concerns.

#### 4.1.3 Pediatrician Density

Primary care pediatrician density had a limited but measurable inverse association with PED use, observed only for CTAS levels 3 and 4*. This suggest that greater outpatient capacity may help reduce moderate, non-urgent visits, though the effect was small. In our counterfactual models, uniformly high pediatrician density accounted for less than 2% of total PED visits, indicating a minor influence compared to proximity or socio-economic factors. This contrasts with previous studies where higher pediatrician supply appeared to reduce PED use in unadjusted analyses, but the effect weakened after adjusting for socio-economic context[33]. No association was observed in unadjusted models, but a modest inverse effect emerged after adjustment, likely reflecting Geneva’s atypical pediatrician distribution, where higher provider densities are found in more socio-economically vulnerable neighborhoods, contrary to patterns in many other high-income settings[34, 35]. Overall, our results suggest that geographic proximity to a pediatrician alone is insufficient to meaningfully curb non-urgent PED use.

### 4.2 Strengths and Limitations

A major strength of this study is the use of a complete, two-year dataset from a public PED in a well-defined urban setting, combined with precise geospatial and sociodemographic data at the neighborhood level. The use of standardized triage levels enabled acuity-specific analyses, and modeling distance as an exponential function allowed for a nuanced characterization of its non-linear effect. The application of fully adjusted mixed-effect models further minimized potential confounding.

However, this study also has several limitations. First, we did not directly assess individual health needs or morbidity, which may partly explain higher PED use in more vulnerable neighborhoods. Although structural and access-related factors were central to our analysis, unmeasured health differences could contribute. Second, the ecological design limits inference at the individual level and introduces potential ecological bias. Finally, the study focused on a single city, which may limit generalizability to other regions with different healthcare systems or population composition. Replication in multicenter studies is warranted to strengthen external validity.

## 5 CONCLUSION AND PERSPECTIVES

This study demonstrates that substantial disparities in PED use exist in a compact, high-income city with universal health coverage. Utilization patterns were shaped not only by need, but also by spatial proximity and neighborhood socio-economic context. Proximity alone accounted up to 15% of visits, particularly for low-acuity cases, while socio-economic vulnerability was associated with even larger disparities. In contrast, pediatrician density had only a modest role. These findings indicate that addressing these disparities may require targeted, locally tailored strategies, such as community outreach in neighborhoods near hospitals, initiatives to build trust and engagement with primary care, and programs to improve health literacy among families.

## 6 DATA SHARING

This study was authorized as quality-of-care study the Geneva Research Ethics Commission (req-2025-00269) and HUG’s institutional board for non-HRA research, but no authorization to share patients’ data was granted.

Data will be made available upon request, conditioned to the approval of a proposal with a signed data access agreement and to the approval of the ethical commission concerned.

## Abbreviation

PED: Pediatric emergency departments
CTAS: Canadian Triage Acuity Scale

## Authors’ contributions

Denis Mongin, Hervé Spechbach, Joachim Marti, Frederic Herler and Johan N. Siebert conceptualized the study.

Denis Mongin, Johan Siebert and Joachim Marti designed the study.

Denis Mongin performed the data collection and curation, carried out the analysis, created the visualisation.

Denis Mongin and Johan Siebert drafted the initial manuscript and critically reviewed and revised the manuscript.

Hervé Spechbach, Joachim Marti and Frederic Herler critically reviewed and revised the manuscript. All authors approved the final manuscript as submitted and agree to be accountable for all aspects of the work

## REFERENCES

1. Calicchio M, Valitutti F, Della Vecchia A, De Anseris AGE, Nazzaro L, Bertrando S, et al. Use and misuse of emergency room for children: features of walk-in consultations and parental motivations in a hospital in Southern Italy. Front Pediatr. 2021;9:674111. Epub 2021/06/26. doi: 10.3389/fped.2021.674111. PubMed PMID: 34169048.

2. Farion KJ, Wright M, Zemek R, Neto G, Karwowska A, Tse S, et al. Understanding low-acuity visits to the pediatric emergency department. PLoS One. 2015;10(6):e0128927. Epub 2015/06/18. doi: 10.1371/journal.pone.0128927. PubMed PMID: 26083338.

3. Ziemnik L, Parker N, Bufi K, Waters K, Almeda J, Stolfi A. Low-acuity pediatric emergency department utilization: caregiver motivations. Pediatr Emerg Care. 2024;40(9):668–73. Epub 2024/03/27. doi: 10.1097/PEC.0000000000003195. PubMed PMID: 38534003.

4. Nicholson E, McDonnell T, De Brun A, Barrett M, Bury G, Collins C, et al. Factors that influence family and parental preferences and decision making for unscheduled pediatric healthcare - systematic review. BMC Health Serv Res. 2020;20(1):663. Epub 2020/07/19. doi: 10.1186/s12913-020-05527-5. PubMed PMID: 32680518.

5. Cairns C, Kang K. National Hospital Ambulatory Medical Care Survey: 2022 Emergency Department Summary Tables. 2022.

6. Jaboyedoff M, Starvaggi C, Suris JC, Kuehni CE, Gehri M, Keitel K. Drivers for low-acuity pediatric emergency department visits in two tertiary hospitals in Switzerland: a cross-sectional, questionnaire-based study. BMC Health Serv Res. 2024;24(1):103. Epub 2024/01/19. doi: 10.1186/s12913-023-10348-3. PubMed PMID: 38238764.

7. Pol A, Biagioli V, Adriani L, Fadda G, Gawronski O, Cirulli L, et al. Non-urgent presentations to the pediatric emergency department: a literature review. Emerg Nurse. 2023;31(5):35–41. Epub 2023/02/03. doi: 10.7748/en.2023.e2154. PubMed PMID: 36727259.

8. Poku BA, Hemingway P. Reducing repeat pediatric emergency department attendance for non-urgent care: a systematic review of the effectiveness of interventions. Emerg Med J. 2019;36(7):435–42. Epub 2019/06/23. doi: 10.1136/emermed-2018-207536. PubMed PMID: 31227526.

9. Morley C, Unwin M, Peterson GM, Stankovich J, Kinsman L. Emergency department crowding: a systematic review of causes, consequences and solutions. PLoS One. 2018;13(8):e0203316. Epub 2018/08/31. doi: 10.1371/journal.pone.0203316. PubMed PMID: 30161242.

10. Kelen GD, Wolfe R, D’Onofrio G, Mills AM, Diercks D, Stern SA, et al. Emergency department crowding: the canary in the health care system. NEJM Catalyst Innovations in Care Delivery. 2021;2(5).

11. Amjad S, Tromburg C, Adesunkanmi M, Mawa J, Mahbub N, Campbell S, et al. Social determinants of health and pediatric emergency department outcomes: a systematic review and meta-analysis of observational studies. Ann Emerg Med. 2024;83(4):291–313. Epub 2023/12/10. doi: 10.1016/j.annemergmed.2023.10.010. PubMed PMID: 38069966.

12. Mathison DJ, Chamberlain JM, Cowan NM, Engstrom RN, Fu LY, Shoo A, et al. Primary care spatial density and nonurgent emergency department utilization: a new methodology for evaluating access to care. Acad Pediatr. 2013;13(3):278–85. Epub 2013/05/18. doi: 10.1016/j.acap.2013.02.006. PubMed PMID: 23680346.

13. Brousseau DC, Hoffmann RG, Nattinger AB, Flores G, Zhang Y, Gorelick M. Quality of primary care and subsequent pediatric emergency department utilization. Pediatrics. 2007;119(6):1131–8. Epub 2007/06/05. doi: 10.1542/peds.2006-3518. PubMed PMID: 17545380.

14. Henneman PL, Garb JL, Capraro GA, Li H, Smithline HA, Wait RB. Geography and travel distance impact emergency department visits. J Emerg Med. 2011;40(3):333–9. Epub 2009/12/17. doi: 10.1016/j.jemermed.2009.08.058. PubMed PMID: 20005663.

15. Kelekar U, Das Gupta D, Theis-Mahon N, Fashingbauer E, Huang B. Distances to emergency departments and non-urgent utilization of medical services: a systematic review. Glob Health Action. 2024;17(1):2353994. Epub 2024/06/03. doi: 10.1080/16549716.2024.2353994. PubMed PMID: 38828477.

16. Bettenhausen JL, Winterer CM, Colvin JD. Health and poverty of rural children: an under-researched and under-resourced vulnerable population. Acad Pediatr. 2021;21(8S):S126–S33. Epub 2021/11/07. doi: 10.1016/j.acap.2021.08.001. PubMed PMID: 34740419.

17. Ramesh T, Yu H. US Pediatric primary care physician workforce in rural areas, 2010 to 2020. JAMA Netw Open. 2023;6(9):e2333467. Epub 2023/09/13. doi: 10.1001/jamanetworkopen.2023.33467. PubMed PMID: 37703020.

18. Warren DW, Jarvis A, LeBlanc L, Gravel J. CTAS National Working Group, Canadian Association of Emergency, Physicians National Emergency Nurses. Revisions to the Canadian Triage and Acuity Scale pediatric guidelines (PaedCTAS). CJEM. 2008;10(3):224–43. Epub 2008/11/21. PubMed PMID: 19019273.

19. Geneva Cantonal Office of Statistics. Cantonal Statistics. In: Department of finance, human resources, and external affairs, editor. Geneva: OCSTAT; 2025. Available from: https://statistique.ge.ch/tel/publications/2025/informations_statistiques/autres_themes/is_population_02_2025.pdf

20. Li H, Calder CA, Cressie N. Beyond Moran’s I: testing for spatial dependence based on the spatial autoregressive model. Geographical analysis. 2007;39(4):357–75.

21. Efron B. Regression and ANOVA with zero-one data: Measures of residual variation. Journal of the American Statistical Association. 1978;73(361):113–21.

22. Marjorie F, Pfarrwaller E, Rozsnyai Z, Streit S, Zurkinden E, Rodondi N, et al. Primary care physicians in Switzerland: state of play and future directions. 2023. Available from: https://www.unifr.ch/med/imf/de/assets/public/images/Research/Primary%20care%20physicians%20in%20Switzerland%20State%20of%20play%20and%20future%20directions%20SAFMED%202023%20(1).pdf

23. Rudge GM, Mohammed MA, Fillingham SC, Girling A, Sidhu K, Stevens AJ. The combined influence of distance and neighbourhood deprivation on emergency department attendance in a large English population: a retrospective database study. PLoS One. 2013;8(7):e67943. Epub 2013/07/23. doi: 10.1371/journal.pone.0067943. PubMed PMID: 23874473.

24. Ludwick A, Fu R, Warden C, Lowe RA. Distances to emergency department and to primary care provider’s office affect emergency department use in children. Acad Emerg Med. 2009;16(5):411–7. Epub 2009/04/25. doi: 10.1111/j.1553-2712.2009.00395.x. PubMed PMID: 19388919.

25. AlSaeed H, Sucha E, Bhatt M, Mitsakakis N, Bresee N, Bechard M. Rates of pediatric emergency department visits vary according to neighborhood marginalization in Ottawa, Canada. CJEM. 2024;26(2):119–27. Epub 2023/12/20. doi: 10.1007/s43678-023-00625-9. PubMed PMID: 38117415.

26. Machuel P, Nafilyan V, Gethings O. Inequalities in Accident and Emergency department attendance, England: March 2021 to March 20222023. Available from: https://www.ons.gov.uk/peoplepopulationandcommunity/healthandsocialcare/healthcaresystem/articles/inequalitiesinaccidentandemergencydepartmentattendanceengland/march2021tomarch2022#cite-this-article.

27. Jensen KV, Morrison A, Ma K, Alqurashi W, Erickson T, Curran J, et al. Low caregiver health literacy is associated with non-urgent pediatric emergency department use. CJEM. 2025;27(1):17–26. Epub 2024/09/27. doi: 10.1007/s43678-024-00771-8. PubMed PMID: 39331337.

28. Morrison AK, Schapira MM, Gorelick MH, Hoffmann RG, Brousseau DC. Low caregiver health literacy is associated with higher pediatric emergency department use and nonurgent visits. Acad Pediatr. 2014;14(3):309–14. Epub 2014/04/29. doi: 10.1016/j.acap.2014.01.004. PubMed PMID: 24767784.

29. Acquadro-Pacera G, Valente M, Facci G, Molla Kiros B, Della Corte F, Barone-Adesi F, et al. Exploring differences in the utilization of the emergency department between migrant and non-migrant populations: a systematic review. BMC Public Health. 2024;24(1):963. Epub 2024/04/06. doi: 10.1186/s12889-024-18472-3. PubMed PMID: 38580984.

30. Brousseau DC, Bergholte J, Gorelick MH. The effect of prior interactions with a primary care provider on nonurgent pediatric emergency department use. Arch Pediatr Adolesc Med. 2004;158(1):78–82. Epub 2004/01/07. doi: 10.1001/archpedi.158.1.78. PubMed PMID: 14706963.

31. Christakis DA, Mell L, Koepsell TD, Zimmerman FJ, Connell FA. Association of lower continuity of care with greater risk of emergency department use and hospitalization in children. Pediatrics. 2001;107(3):524–9. Epub 2001/03/07. doi: 10.1542/peds.107.3.524. PubMed PMID: 11230593.

32. Durand AC, Palazzolo S, Tanti-Hardouin N, Gerbeaux P, Sambuc R, Gentile S. Nonurgent patients in emergency departments: rational or irresponsible consumers? Perceptions of professionals and patients. BMC Res Notes. 2012;5:525. Epub 2012/09/26. doi: 10.1186/1756-0500-5-525. PubMed PMID: 23006316.

33. Michelson KA, Cushing AM, Bucholz EM. Association of county-level availability of pediatricians with emergency department visits. Pediatr Emerg Care. 2022;38(2):e953–e7. Epub 2021/07/21. doi: 10.1097/PEC.0000000000002502. PubMed PMID: 34282091.

34. Freed GL, Nahra TA, Wheeler JR, Research Advisory Committee of the American Board of Pediatrics. Relation of per capita income and gross domestic product to the supply and distribution of pediatricians in the United States. J Pediatr. 2004;144(6):723–8. Epub 2004/06/12. doi: 10.1016/j.jpeds.2004.02.043. PubMed PMID: 15192616.

35. Mancheron A, Vincent-Cassy C, Guedj R, Chappuy H, De Groc T, Duval Arnould M, et al. Association between socio-economic status and nonurgent presentations to pediatric emergency departments: a retrospective study. Eur J Emerg Med. 2025. Epub 2025/01/24. doi: 10.1097/MEJ.0000000000001217. PubMed PMID: 39854298.

