## Supplementary material for "Driving Factors in Pediatric Emergency Department Use: an Ecological Retrospective Study"

#### Supplementary for methods

Full model with interaction term:

$$Y_{i,j}\sim A\exp\left( \frac{-\ln\left( 2 \right)\times dist_{i,j}}{\lambda} \right)+{{(b}_{0}+b}_{j})+c\times NSVI_{i,j}+Ac\times NSVI_{i,j}\times exp \left( \frac{-ln \left( 2 \right)\times dist_{i,j}}{\lambda} \right)+d\times(PEDdensity_{i,j}-1)+\epsilon_{i,j}$$

Ac is the coefficient measuring the interaction between NSVI and the non-linear distance effect.

#### Supplementary tables and figures:

**Supplementary Table 1**: Composition of the neighbourhood socio-economic vulnerability index (NSVI). Each variable is computed for the geographical neighbourhood.

| **Variable** | **Threshold defining a low socio-economic level in the neighbourhood** |
| --- | --- |
| Proportion of housing allowance | Above the fourth quartile of the overall proportion of housing allowance |
| Median income | Below the first quartile of the overall distribution of median income |
| share of low income^1^ | Above the fourth quartile of the overall share of low income |
| share of students coming from a modest family_2_ | Above the fourth quartile of the overall share of student coming from a modest family |
| share of active persons registered to the unemployment office | Above the fourth quartile of the overall share of active persons registered to the unemployment office |
| share of person perceiving social subsidies | Above the fourth quartile of the overall person perceiving social subsidies |

^1^Low income is considered as person having an income within the first quartile of the canton income distribution.

^2^students coming from a modest family are student with their parent designed by the professional category coded as “blue-collar worker” or “other” in the database of education.

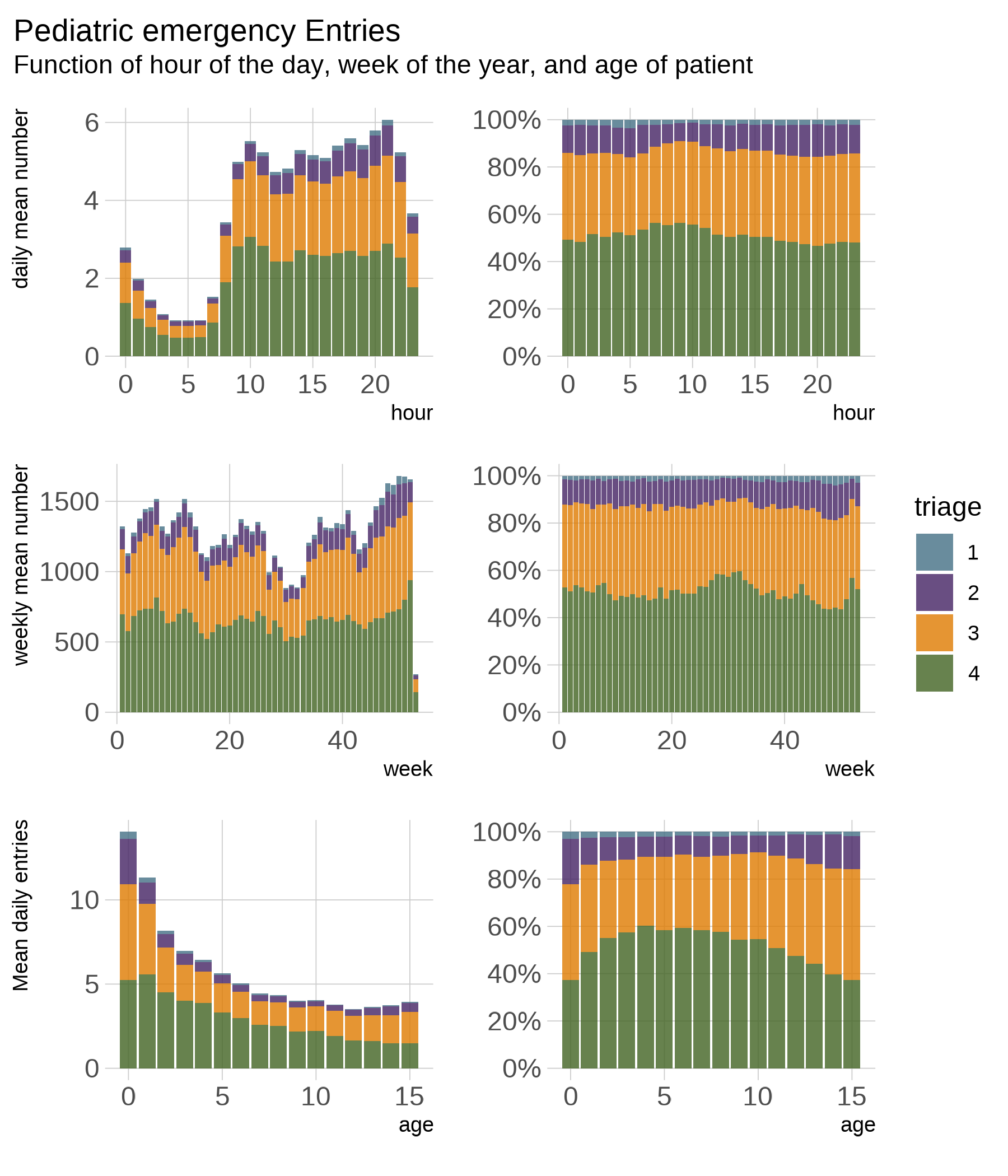

**Supplementary Figure 1**: Number of PED entries as a function of the hour of the day (top row), of the day of the year (middle row), and of the age of the children (bottom row), stratified by Canadian Triage Acuity Scale (CTAS) level (2: emergent, 3: urgent, 4*: less urgent (4) and non-urgent (5)). Left column: absolute numbers; right column: proportion.

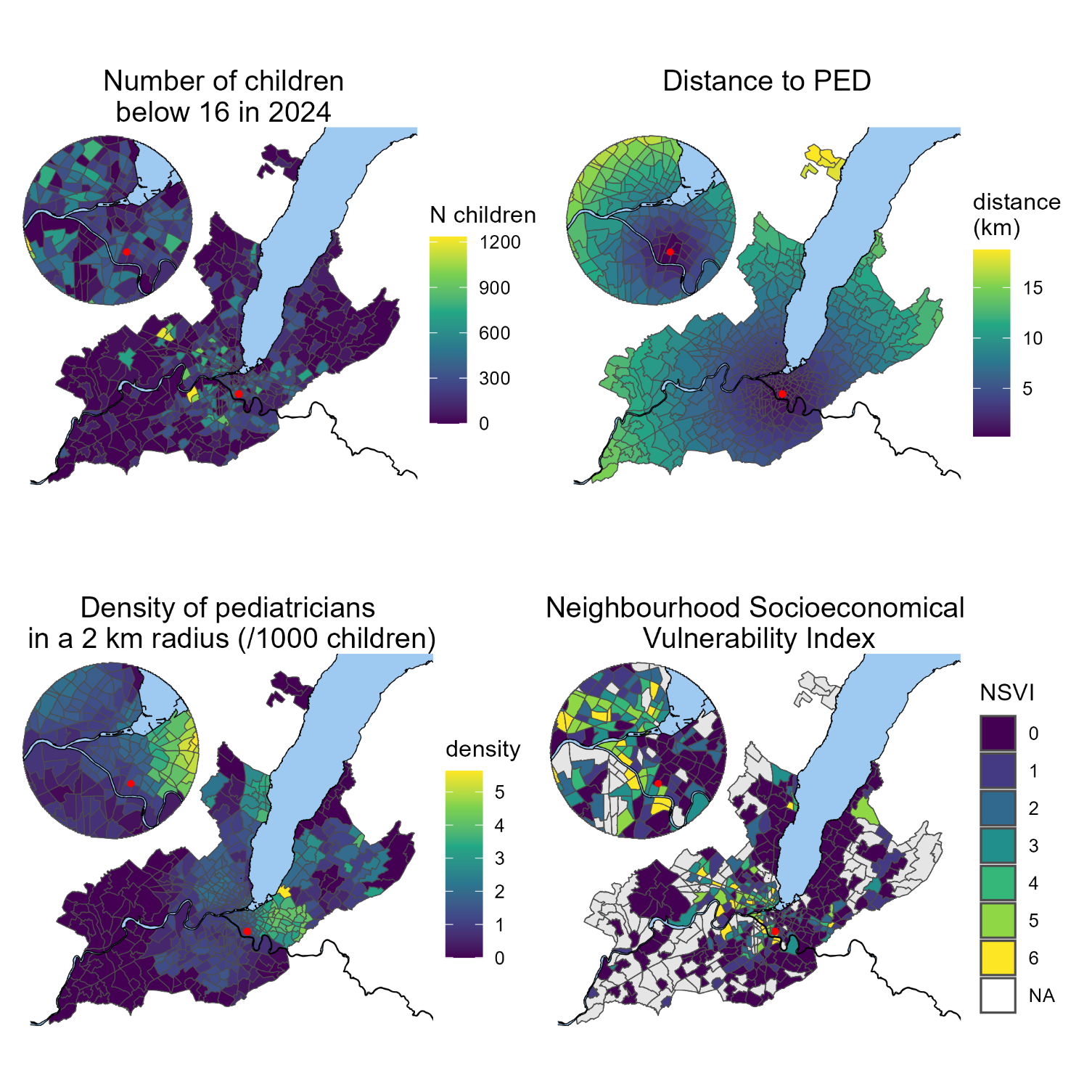

**Supplementary figure 2**: spatial distribution in the state of Geneva (per neighbourhood) of the Number of children, distance to the Pediatric Emergency department, the density of pediatricians in a 2 km radius, and of the socioeconomical vulnerability index.

Sensibility analysis: patients aged 0-5 years

| CTAS  Coeff | 1 | 2 | 3 | 4 |
| --- | --- | --- | --- | --- |
| Spatial effects: maximum amplitude (A) | -1.3 [-5.6; 3] (-64%) | 6.7 [4.3; 9.1]*** (65.6%) | 19 [ 14; 24]*** (78.1%) | 33 [ 26; 39]*** (104%) |
| characteristic length (λ , in km) | 0.013 [-0.073; 0.1] | 2 [0.23; 3.8]* | 2.7 [0.76; 4.7]** | 2.7 [1.2; 4.2]*** |
| pediatrician density (c) | -0.15 [-0.87; 0.56] (-7.5%) | -1.5 [-3.3; 0.24] (-14.9%) | -2.3 [-5.7; 0.97] (-9.6%) | -4.7 [ -9; -0.38]* (-14.9%) |
| socioeconomic effect (d) | 0.07 [-0.053; 0.19] (20.4%) | 0.68 [0.38; 0.98]*** (40.2%) | 1.9 [1.4; 2.5]*** (48%) | 2.3 [1.6; 3]*** (43.2%) |
| baseline incidence (b) | 1.9 [1.4; 2.4]*** (91%) | 5.9 [3.7; 8.1]*** (58.2%) | 12 [ 6; 17]*** (47.7%) | 11 [4.1; 18]** (35.7%) |
| Moran Correlation index of the residuals | -0.017 | 0.032 | -0.0072 | -0.072 |
| Median incidence | 2.1 | 10 | 24 | 31 |

**Supplementary table 2**: Results of the multivariable regression modeling the incidence of PED use for children between 0 and 5 years during 2023-2024 as a function of distance to the PED, neighborhood socio-economic vulnerability, and pediatrician density. $A$ and $\lambda$ represent the amplitude and characteristic length of the exponential spatial decay, respectively. $c$ is the coefficient for pediatrician density (expressed as the number of pediatricians per 1000 children), $d$ is the coefficient for Neighbourhood Socio-Economic Vulnerability Index (NSVI), and b denotes the baseline incidence. The percentage in parenthesis represents the marginal variation induced by the variable compared to the mean incidence.

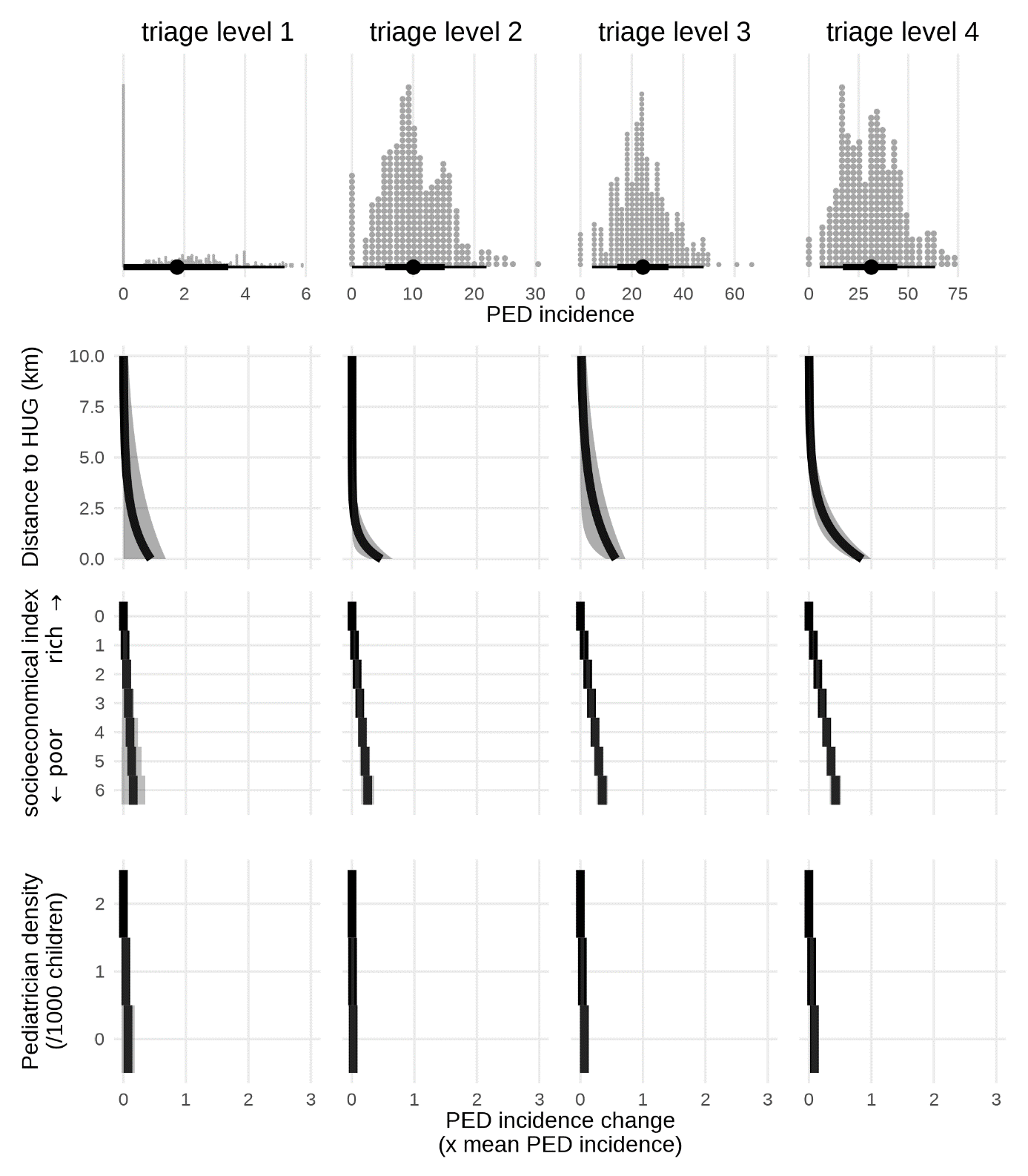

**Supplementary Figure 3**: effect of the different exposure of interest on the incidence of PED use for children ages 0 to 5 years, stratified by triage level (columns). The top row represents the distribution of the neighbourhood incidences. The three bottom rows represent the PED use incidence variation induced by the distance to PED, the neighbourhood socio-economical index, and the paediatrician density, expressed as a factor of the mean PED use incidence.

### Sensibility analysis: patients aged 6-15 years

| CTAS  coeff | 1 | 2 | 3 | 4 |
| --- | --- | --- | --- | --- |
| Spatial effects: maximum amplitude (A) | 0.9 [0.52; 1.3]*** (165.6%) | 3.4 [1.6; 5.2]*** (95%) | 11 [7.3; 14]*** (86.5%) | 23 [ 18; 28]*** (143.5%) |
| characteristic length (λ , in km) | 1.9 [-0.0042; 3.9] | 0.61 [0.092; 1.1]* | 1.6 [0.57; 2.6]** | 1.1 [0.72; 1.6]*** |
| pediatrician density (c) | -0.36 [-0.89; 0.17] (-65.7%) | -0.63 [-2.2; 0.95] (-17.9%) | -3.6 [-7.8; 0.54] (-28.9%) | -6 [-11; -0.69]* (-37.4%) |
| socioeconomic effect (d) | 0.066 [0.016; 0.12]** (72.6%) | 0.24 [0.089; 0.38]** (40.2%) | 1.1 [0.81; 1.5]*** (55.2%) | 1.9 [1.5; 2.4]*** (71.4%) |
| baseline incidence (b) | -0.12 [-0.66; 0.43] (-21.5%) | 2.4 [ 1; 3.7]*** (66.9%) | 4.9 [0.83; 9.1]* (39.6%) | 3.9 [-1.1; 8.8] (23.9%) |
| Moran Correlation index of the residuals | -0.002 | 0.1 | 0.0018 | 0.028 |
| Median incidence | 0.54 | 3.5 | 12 | 16 |

Supplementary table 3: Results of the multivariable regression modeling the incidence of PED use for children between 6 and 15 years during 2023-2024 as a function of distance to the PED, neighborhood socio-economic vulnerability, and pediatrician density. $A$ and $\lambda$ represent the amplitude and characteristic length of the exponential spatial decay, respectively. $c$ is the coefficient for pediatrician density (expressed as the number of pediatricians per 1000 children), $d$ is the coefficient for Neighbourhood Socio-Economic Vulnerability Index (NSVI), and b denotes the baseline incidence. The percentage in parenthesis represents the marginal variation induced by the variable compared to the mean incidence.

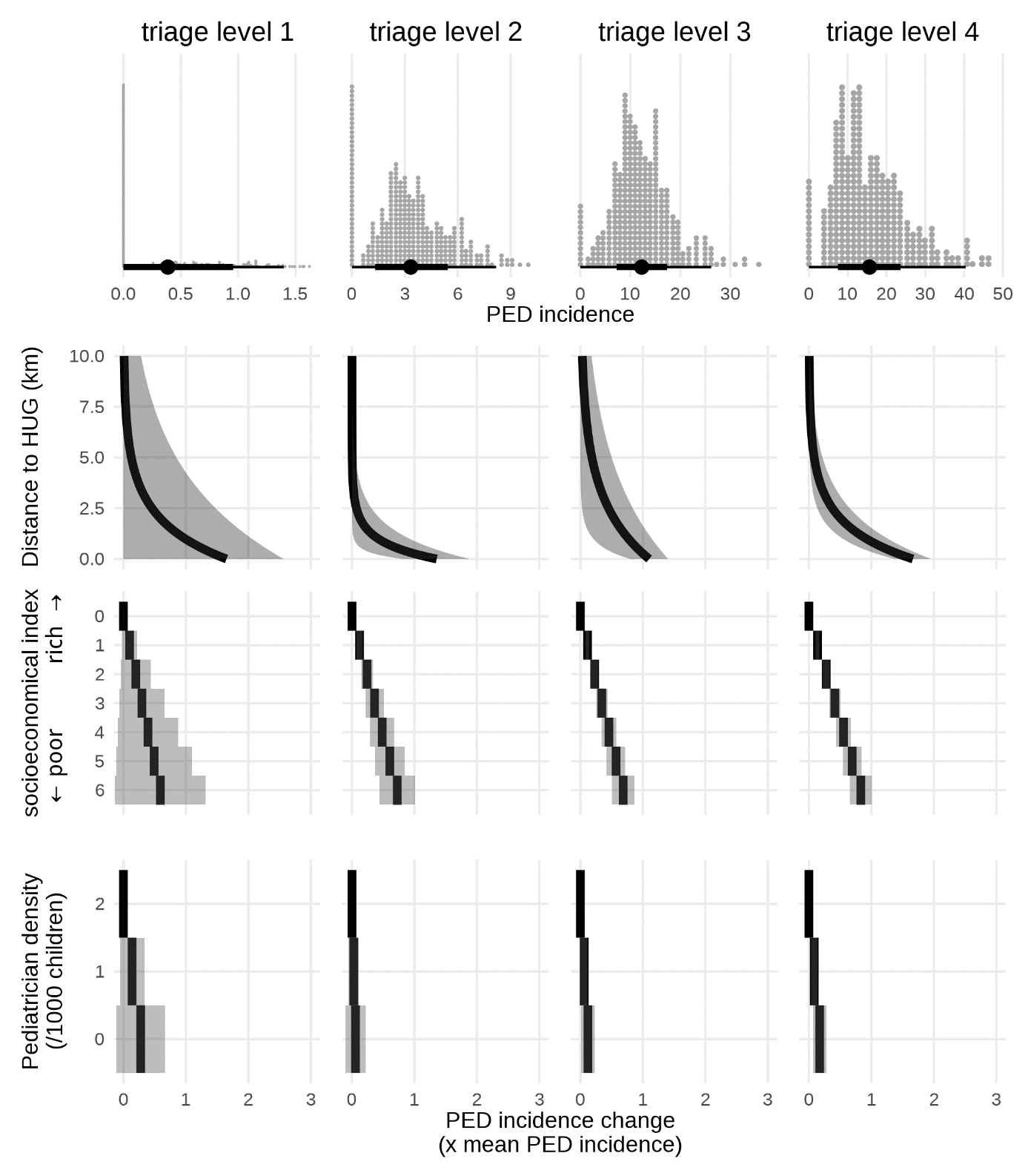

Supplementary Figure 4: effect of the different exposure of interest on the incidence of PED use for children ages 6 to 15 years, stratified by triage level (columns). The top row represents the distribution of the neighbourhood incidences. The three bottom rows represent the PED use incidence variation induced by the distance to PED, the neighbourhood socio-economical index, and the paediatrician density, expressed as a factor of the mean PED use incidence.

##### Sensibility analysis: during school hours (Monday – Friday, 7h – 18h)

| CTAS  coeff | 1 | 2 | 3 | 4 |
| --- | --- | --- | --- | --- |
| Spatial effects: maximum amplitude (A) | 0.51 [0.13; 0.89]** (118.6%) | 2.2 [1.1; 3.3]*** (80.8%) | 8.9 [6.6; 11]*** (105%) | 21 [ 17; 25]*** (193.2%) |
| characteristic length (λ , in km) | 3.5 [-3.6; 11] | 0.93 [0.14; 1.7]* | 1.8 [0.77; 2.8]*** | 0.92 [0.62; 1.2]*** |
| pediatrician density (c) | -0.39 [ -1; 0.24] (-90.7%) | -0.77 [-2.5; 0.99] (-28.4%) | -4.5 [-9.1; 0.066] (-53.3%) | -8.6 [-15; -2]* (-78.9%) |
| socioeconomic effect (d) | 0.019 [-0.02; 0.058] (25.8%) | 0.27 [0.16; 0.37]*** (58.8%) | 0.95 [0.67; 1.2]*** (67.2%) | 1.6 [1.3; 2]*** (90%) |
| baseline incidence (b) | -0.17 [-0.92; 0.57] (-40.3%) | 1.3 [-0.27; 2.9] (48.9%) | 0.58 [-3.9; 5] (6.9%) | -2.2 [-8.2; 3.8] (-20.5%) |
| Moran Correlation index of the residuals | -0.0091 | -0.011 | 0.063 | -0.012 |
| Median incidence | 0.43 | 2.7 | 8.4 | 11 |

Supplementary table 4: Results of the multivariable regression modeling the incidence of PED use between 7h and 18h during 2023-2024 as a function of distance to the PED, neighborhood socio-economic vulnerability, and pediatrician density. $A$ and $\lambda$ represent the amplitude and characteristic length of the exponential spatial decay, respectively. $c$ is the coefficient for pediatrician density (expressed as the number of pediatricians per 1000 children), $d$ is the coefficient for Neighbourhood Socio-Economic Vulnerability Index (NSVI), and b denotes the baseline incidence. The percentage in parenthesis represents the marginal variation induced by the variable compared to the mean incidence.

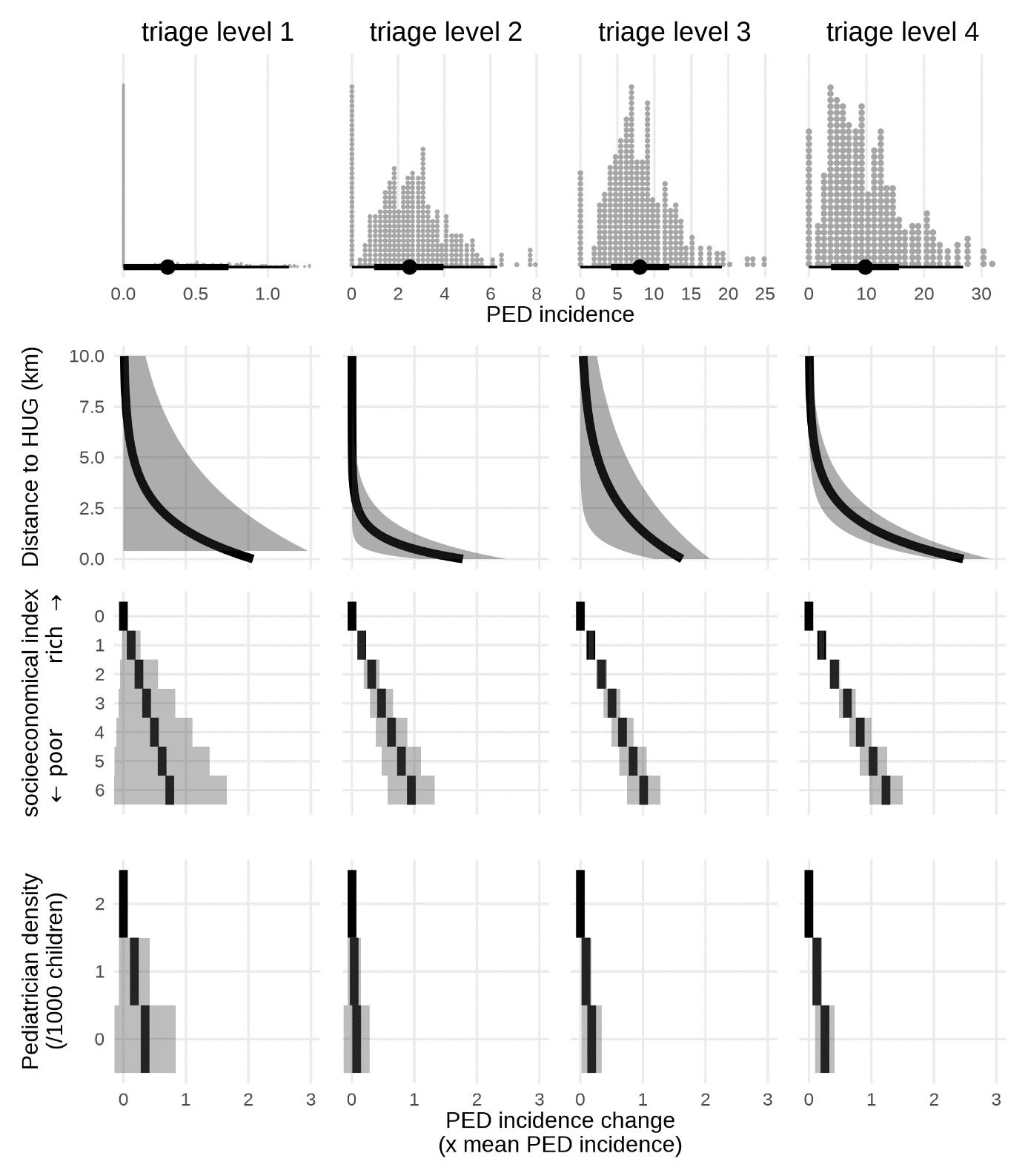

Supplementary Figure 5: effect of the different exposure of interest on the incidence of PED use during the week school hours (7h-18h), stratified by triage level (columns). The top row represents the distribution of the neighbourhood incidences. The three bottom rows represent the PED use incidence variation induced by the distance to PED, the neighbourhood socio-economical index, and the paediatrician density, expressed as a factor of the mean PED use incidence.

##### Sensibility analysis: during the week outside of school hours

| CTAS  coeff | 1 | 2 | 3 | 4 |
| --- | --- | --- | --- | --- |
| Spatial effects: maximum amplitude (A) | 0.31 [-0.16; 0.78] (84.1%) | 1.5 [0.57; 2.4]** (69.7%) | 4.2 [2.2; 6.3]*** (73.4%) | 8.5 [6.6; 10]*** (118.8%) |
| characteristic length (λ , in km) | 0.66 [-0.92; 2.2] | 1.2 [-0.087; 2.4] | 3.6 [-1.2; 8.3] | 2.9 [1.2; 4.7]** |
| pediatrician density (c) | -0.27 [-0.93; 0.38] (-73.9%) | -0.58 [-2.1; 0.94] (-27.5%) | -2 [-5.3; 1.2] (-35.4%) | -4.2 [-7.8; -0.67]* (-59.5%) |
| socioeconomic effect (d) | 0.017 [-0.022; 0.057] (28.2%) | 0.12 [0.034; 0.21]** (34.2%) | 0.47 [0.27; 0.67]*** (49.2%) | 0.73 [0.51; 0.95]*** (61.8%) |
| baseline incidence (b) | 0.08 [-0.5; 0.66] (21.5%) | 1.2 [-0.26; 2.6] (54.8%) | 1.3 [-2.7; 5.2] (21.9%) | -1.2 [-5.1; 2.7] (-16.8%) |
| Moran Correlation index of the residuals | -0.037 | -0.021 | -0.072 | -0.0081 |
| Median incidence | 0.37 | 2.1 | 5.7 | 7.1 |

Supplementary table 4: Results of the multivariable regression modeling the incidence of PED use for children between 19h and 06h during 2023-2024during 2023-2024 as a function of distance to the PED, neighborhood socio-economic vulnerability, and pediatrician density. $A$ and $\lambda$ represent the amplitude and characteristic length of the exponential spatial decay, respectively. $c$ is the coefficient for pediatrician density (expressed as the number of pediatricians per 1000 children), $d$ is the coefficient for Neighbourhood Socio-Economic Vulnerability Index (NSVI), and b denotes the baseline incidence. The percentage in parenthesis represents the marginal variation induced by the variable compared to the mean incidence.

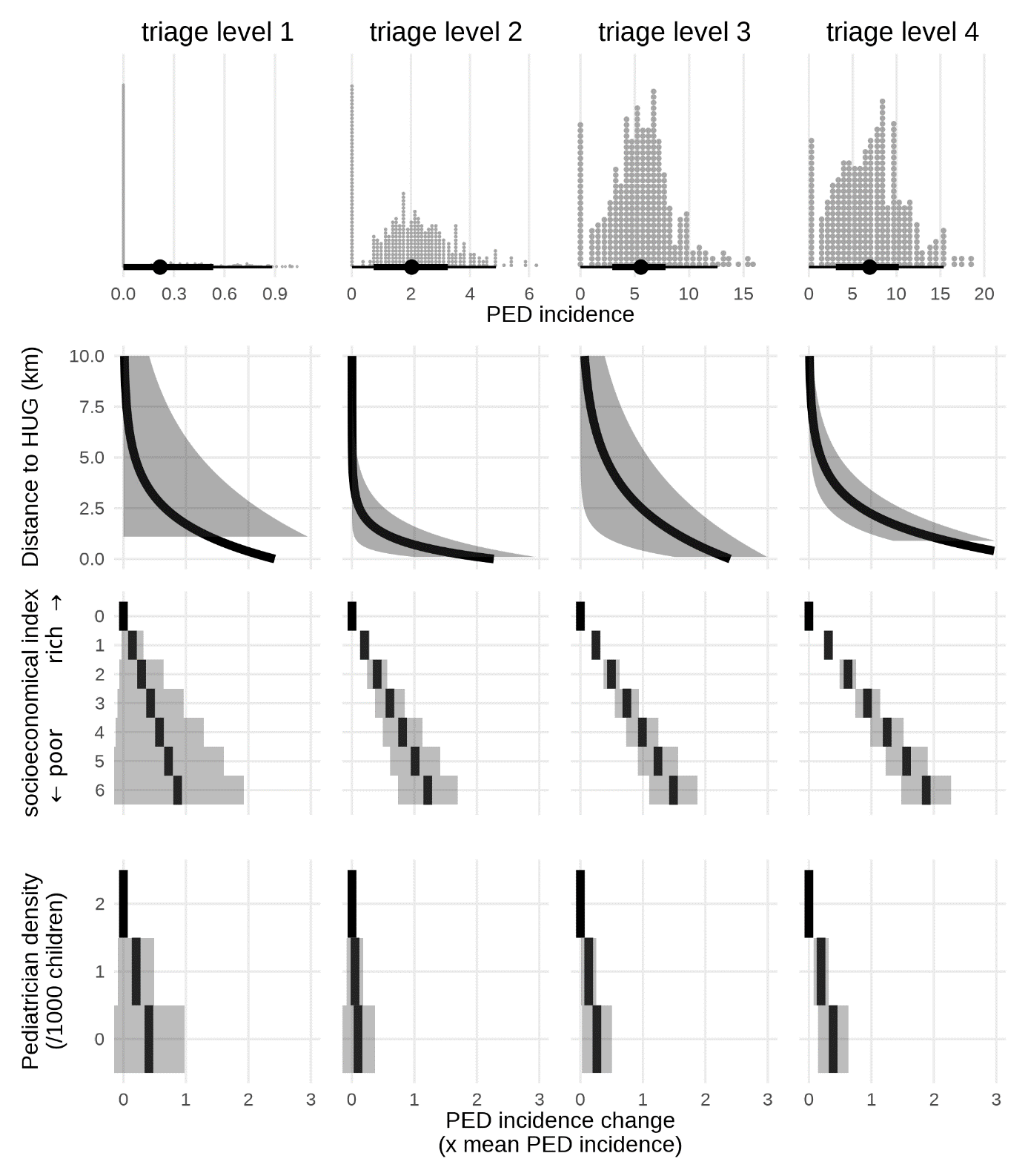

Supplementary Figure 6: effect of the different exposure of interest on the incidence of PED use during the week outside school hours (19h-06h), stratified by triage level (columns). The top row represents the distribution of the neighbourhood incidences. The three bottom rows represent the PED use incidence variation induced by the distance to PED, the neighbourhood socio-economical index, and the paediatrician density, expressed as a factor of the mean PED use incidence.

##### Sensibility analysis: during weekends

| CTAS  coeff | 1 | 2 | 3 | 4 |
| --- | --- | --- | --- | --- |
| Spatial effects: maximum amplitude (A) | 0.36 [0.072; 0.65]* (108.5%) | 1.9 [0.95; 2.9]*** (103.7%) | 7.9 [0.54; 15]* (134.3%) | 10 [7.7; 13]*** (118.4%) |
| characteristic length (λ , in km) | 2.5 [ -3; 8] | 0.83 [0.13; 1.5]* | 7.3 [ -5; 19] | 2.1 [0.88; 3.4]*** |
| pediatrician density (c) | -0.26 [-0.87; 0.35] (-78.2%) | -0.85 [-2.3; 0.64] (-46.1%) | -5.1 [-8.4; -1.8]** (-85.8%) | -6.7 [-12; -1.7]** (-78.6%) |
| socioeconomic effect (d) | 0.027 [-0.011; 0.065] (48.6%) | 0.12 [0.026; 0.21]* (37.8%) | 0.47 [0.27; 0.67]*** (48%) | 1 [0.73; 1.3]*** (72.6%) |
| baseline incidence (b) | -0.069 [-0.7; 0.57] (-20.6%) | 0.68 [-0.66; 2] (37.1%) | -4.5 [-13; 4.4] (-75.6%) | -2.2 [-7.2; 2.9] (-25.4%) |
| Moran Correlation index of the residuals | -0.056 | 0.062 | 0.052 | 0.047 |
| Median incidence | 0.33 | 1.8 | 5.9 | 8.6 |

Supplementary table 6: Results of the multivariable regression modeling the incidence of PED use for children for 2023-2024 weekends (Saturday and Sunday) as a function of distance to the PED, neighborhood socio-economic vulnerability, and pediatrician density. $A$ and $\lambda$ represent the amplitude and characteristic length of the exponential spatial decay, respectively. $c$ is the coefficient for pediatrician density (expressed as the number of pediatricians per 1000 children), $d$ is the coefficient for Neighbourhood Socio-Economic Vulnerability Index (NSVI), and b denotes the baseline incidence. The percentage in parenthesis represents the marginal variation induced by the variable compared to the mean incidence.

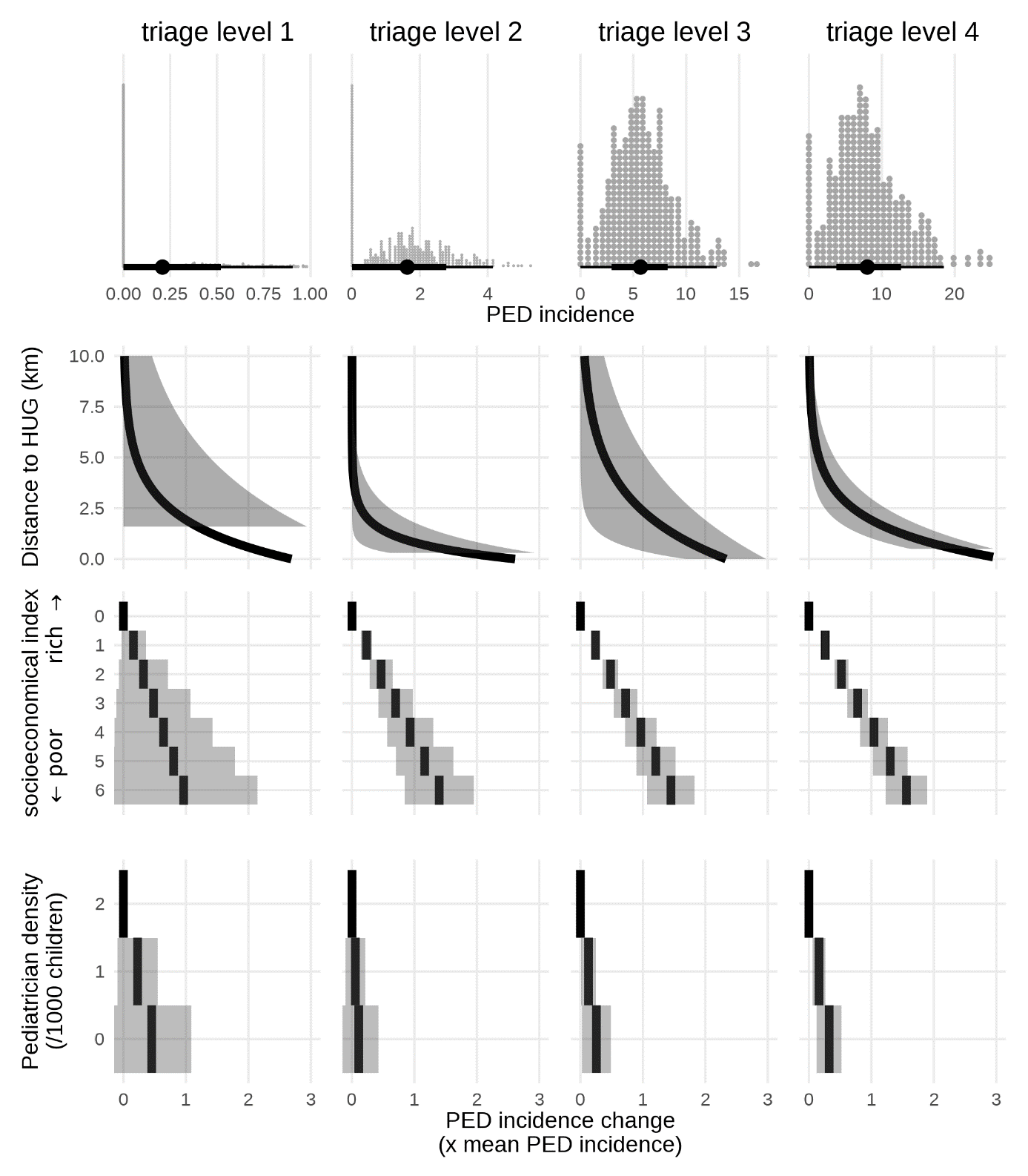

Supplementary Figure 7: effect of the different exposure of interest on the incidence of PED use during the weekends, stratified by triage level (columns). The top row represents the distribution of the neighbourhood incidences. The three bottom rows represent the PED use incidence variation induced by the distance to PED, the neighbourhood socio-economical index, and the paediatrician density, expressed as a factor of the mean PED use incidence.
